# Covid-19 vs BCG: Statistical Significance Analysis

**DOI:** 10.1101/2020.06.08.20125542

**Authors:** Serge Dolgikh

**Affiliations:** National Aviation University

**Keywords:** Infectious epidemiology, statistical analysis, Covid-19

## Abstract

We present an updated time-adjusted dataset and conclusions at Covid-19 Time Zero + 5 month (04.06.2020). The conclusions of the original analysis reviewed and mostly maintained at this time point. With the data accumulated to date a statistical significance of the BCG immunization correlation hypothesis is evaluated with the conclusion that it has achieved the level of confidence. Several specific cases are discussed with respect to the induced immunity hypothesis.

## 1 Data

### 1.1 Terminology

Covid-19 global Time Zero (TZ) was defined in [1] as 31.12.2020: Along with the global Time Zero was defined local Time Zero (LTZ) indicating the time of arrival of the epidemics in the given locality. It can be sensibly defined as the date of the first confirmed case in the area.

The impact of the epidemics is measured by Covid-19 caused mortality per 1 M capita:

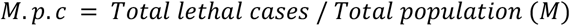

It is believed that this parameter is a more current and accurate measure of the epidemics impact than the number of cases that strongly depends on the testing practice, on the assumption that policies and protocols in the selected administration allow more accurate identification of cause and reporting.

### 1.2 Case Data

The following datasets were compiled from publicly available data:

1. An updated current time snapshot of selected jurisdictions dataset as of approximately, TZ + 5 m (04.06.2020).
2. A time-adjusted selected jurisdictions dataset, with data of Wave 1 and Wave 2 cases adjusted by the time of first exposure to Covid-19. Specifically, the dataset is comprised of the Wave 1 cases as of approx. TZ + 4 m and Wave 2 cases as of, approx. TZ+ 5 m i.e. with approximately the same local exposure of approximately 3 months. The significance of this point in the epidemics development timeline is that most exposed jurisdictions can be expected to have achieved the peak of the impact by this point. BCG universal vaccination record is measured in the following bands as defined in [2]:

A: has universal or near-universal BCG coverage

A2: has current BCG coverage with some limitations or qualifications (such as a late start; inconsistencies in application practice, interruptions and other)

B: had BCG vaccination in the past covering significant part of population (> 50%)

B2: UIP was offered for a limited time interval or specific groups

B3: UIP practice inconsistent with the hypothesis of early age induced immunity protection for example, delivered at an older age

C: never had a UIP/BCG

As well, we define groups of cases by the reported impact of Covid-19 on the population, where relative M.p.c. is measured as a ratio of the local M.p.c. to the world’s highest value, at the time of writing, near 2500 / 1 million.

Very Low (VL), relative m.p.c. near 0.001

Low (L): r.m.p.c. below or near 0.01

Medium: (M): r.m.p.c. below or near 0.1

High (H): relative m.p.c. noticeably higher than 0.1

Several common sense criteria were applied such as: certain expectation of reliability and consistency of the reporting jurisdiction; a reasonable level of exposure to Covid-19, e.g. certain minimum number of reported cases; geographical and development level variation.

#### Disclaimers

1. Consistency and reliability of data reported by the national, regional and local health administrations.
2. Alignment in the time of reporting may be an issue due to reporting practices of jurisdictions.
3. Availability, consistency and reliability of historical data and statistics on the administration of immunization programs in the national, regional and so on, jurisdictions can be an issue.

Case datasets can be found in the Appendix. Sources: [3-11].

## 2. Observations

### 2.1 New Developments

New cases of rapid onset added: *Brazil, Mexico*.

High impact cases in Europe and North America including Netherlands; Ireland; USA and Quebec (Canada) appear to be entering the sustained phase or in decline since the last update.

### 2.2 Effective Management Cases

Low epidemics impact, measured by relative M.p.c. in the range of 0.001-0.003 is maintained in this group. New cluster development in Japan appears to have been stabilized since the previous update. All countries in this group have a current BCG UIP except Australia that had it till mid-1980-ies.

### 2.3 BCG UIP Correlation

The hypothesis of a correlation between a universal immunization program with BCG tuberculosis vaccine (BCG UIP) and milder epidemics impact was proposed in [12] and analyzed based on available at the time data in [1]. The conclusions of the original analysis are mostly maintained at this time point:

All countries in the VL category currently have a UIP or had it recently still covering most of age cohorts. The majority of countries in the L category have an ongoing UIP or had it till recently.

No cases from the group C that never had a UIP are found in the groups VL, L with lower impacts of Covid-19, though regional variations are possible (e.g. Prairie provinces, Canada Canada).

Figure 1 illustrates the distribution of cases with respect to BCG UIP record by epidemiological impact measured as logarithm of M.p.c., time adjusted to LTZ + 3 months. Apart from outliers, the bulk of cases falls into the interval [1.5, 9.5]. Three impact groups defined were “Low”, with the impact in the lower half of the range; and “Mid” and “High”, with the impact in the third and the fourth quarter, respectively.

**Fig. 1.**
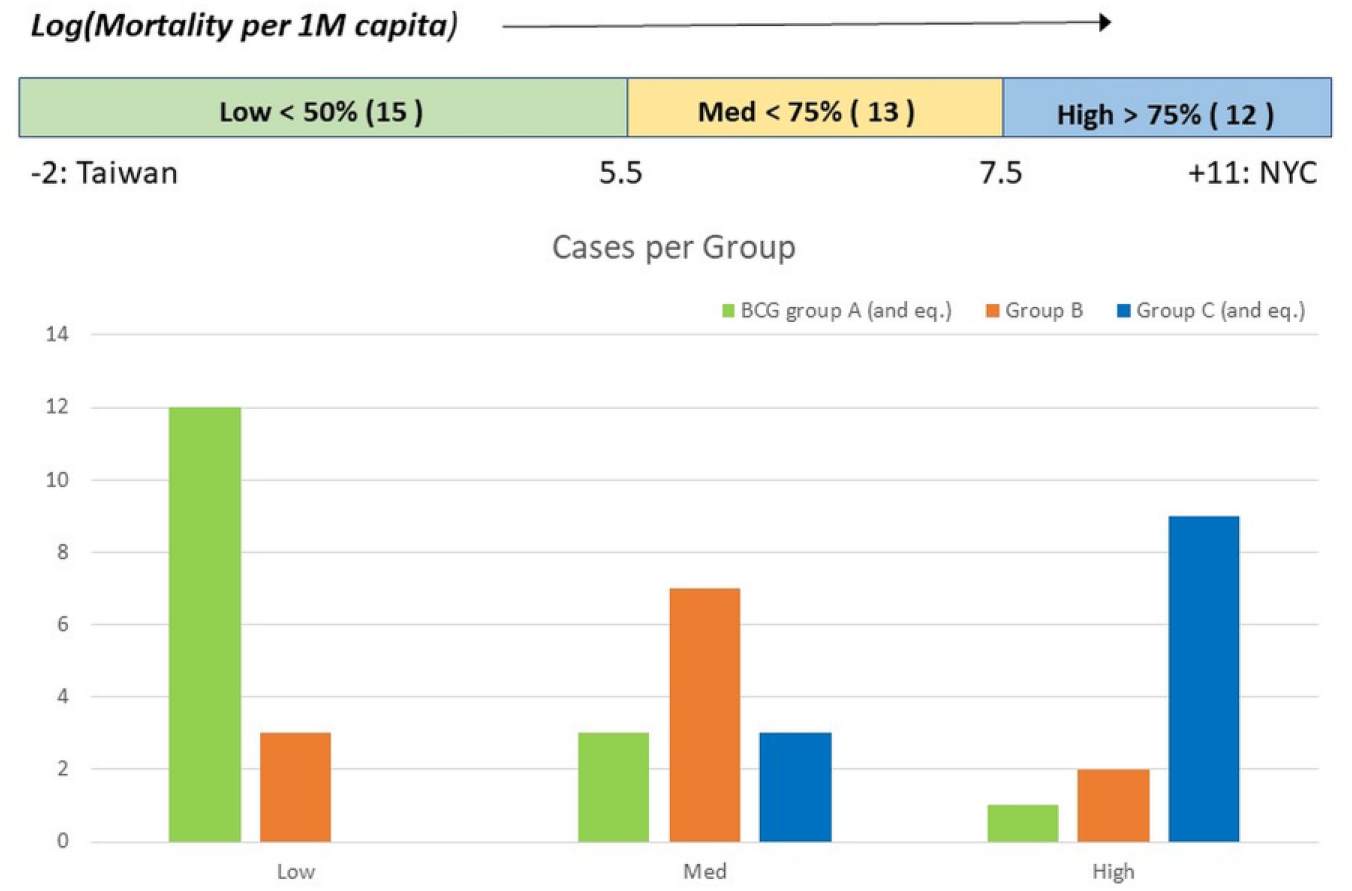
Case distribution by severity of impact.

As can be seen in the diagram, the mean impacts observed in the groups A and equivalent (that included group B cases with a very recent UIP ceased after 2005) and group C and equivalent (that included group B2 cases with a very short UIP such as Spain, 16 years in total and Quebec, Canada 18 years that could not be expected to provide significant protection effect compared to no UIP scenario) clearly tend toward the opposite ends of the impact range.

### 2.4 Cessation of UIP vs. Impact

In the group B, where a BCG immunization program existed but was ceased earlier, a strong correlation can be observed between the time of cessation of the UIP and the severity of Covid-19 impact as shown in the diagram of Fig.2 (the data is time adjusted to LTZ + 3 m):

**Fig. 2.**
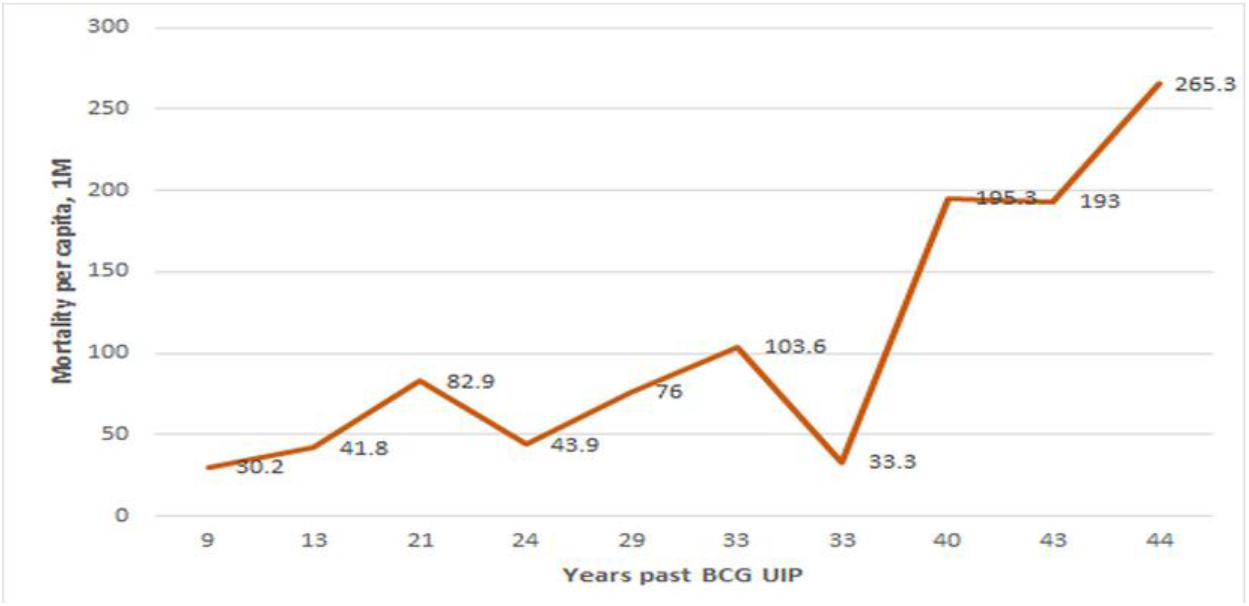
Impact vs. Time past BCG UIP relationship.

**Fig. 3.**
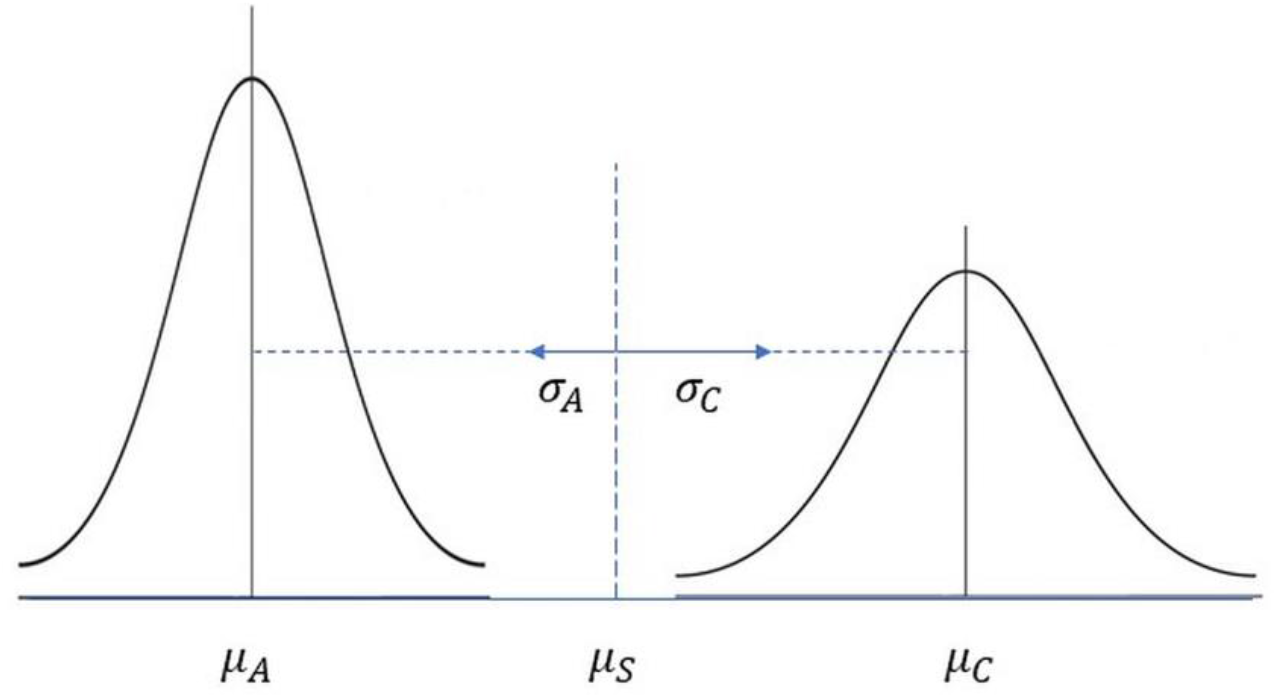
Sample means distributions, BCG groups.

### 2.5 Regional and Age Cohort Variation Analysis

In certain jurisdictions significant regional variability in administration of BCG vaccination can be noted, providing further information relevant to the correlation hypothesis.

#### Portugal – Spain

Portugal: group A2 (current BCG UIP, late start 1965)

Spain: group B3, C-equivalent (short UIP duration, 1965 – 1981)

The observation of a correlation in the original analysis is strongly maintained at LTZ+3: Portugal, M.p.c *139.4* vs. Spain, *544.5*.

#### Northern Europe

Adjusted to the same time of local exposure (LTZ + 3 m), the four cases of Northern Europe show strong correlation between Covid-19 impact and the time of cessation of BCG UIP (Table 3).

**Table 1.**
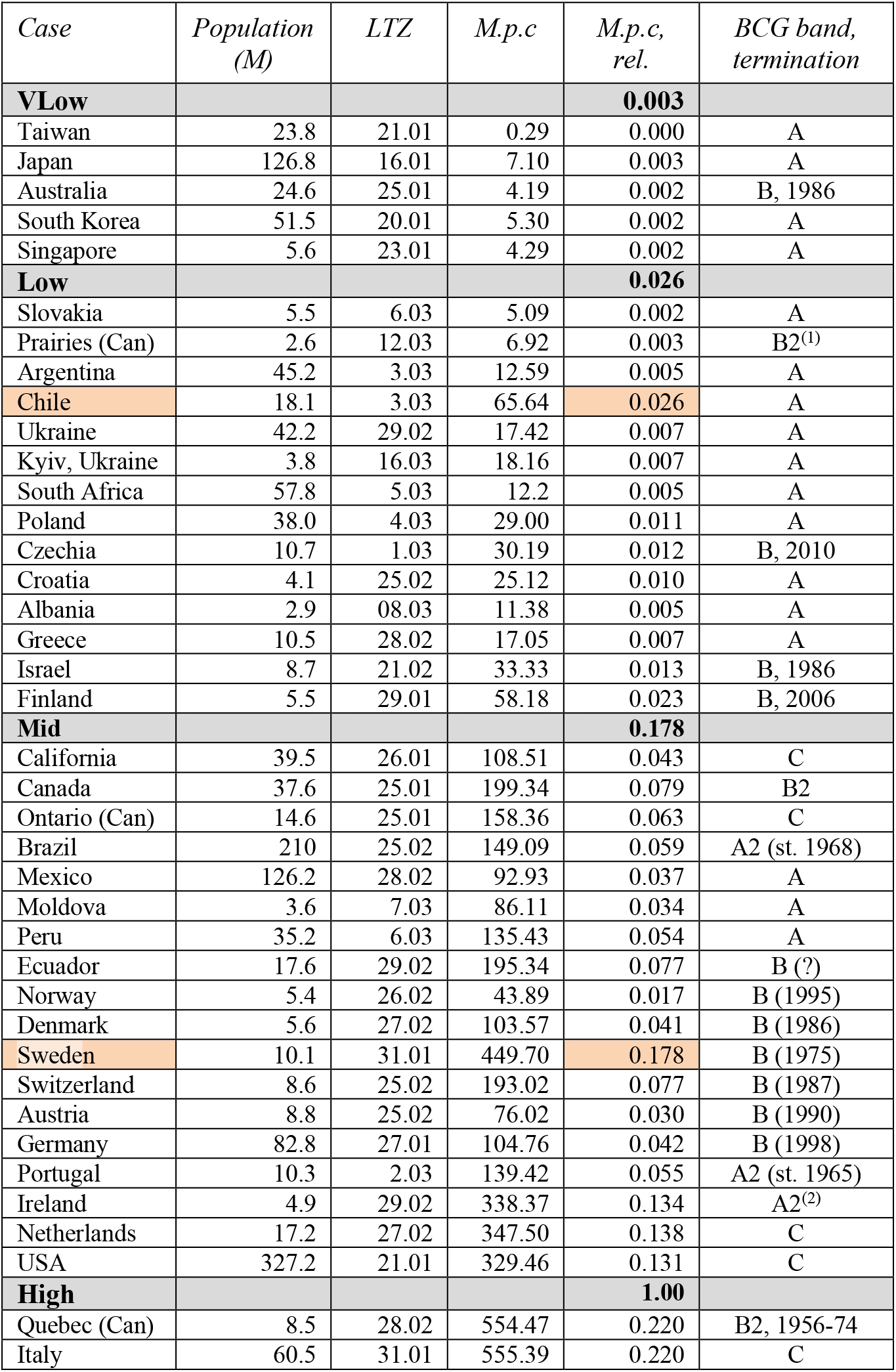
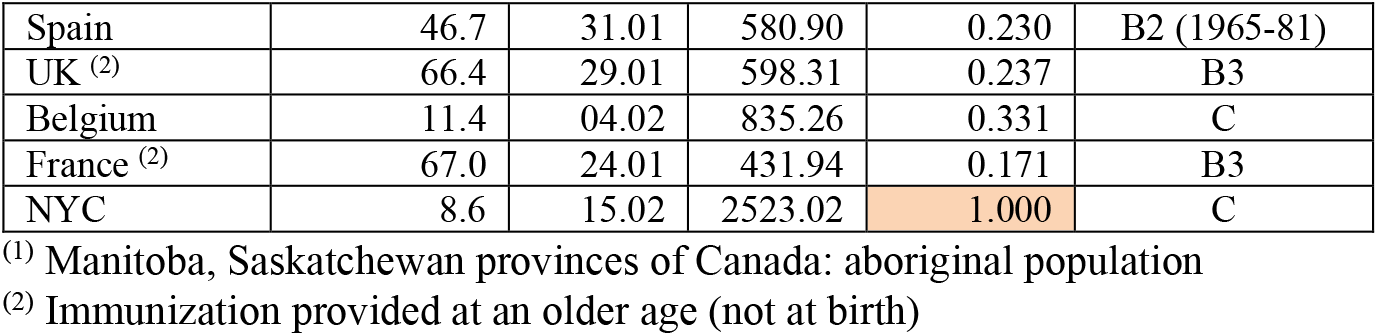
Current Dataset (updated 04.06.2020)

**Table 2.**
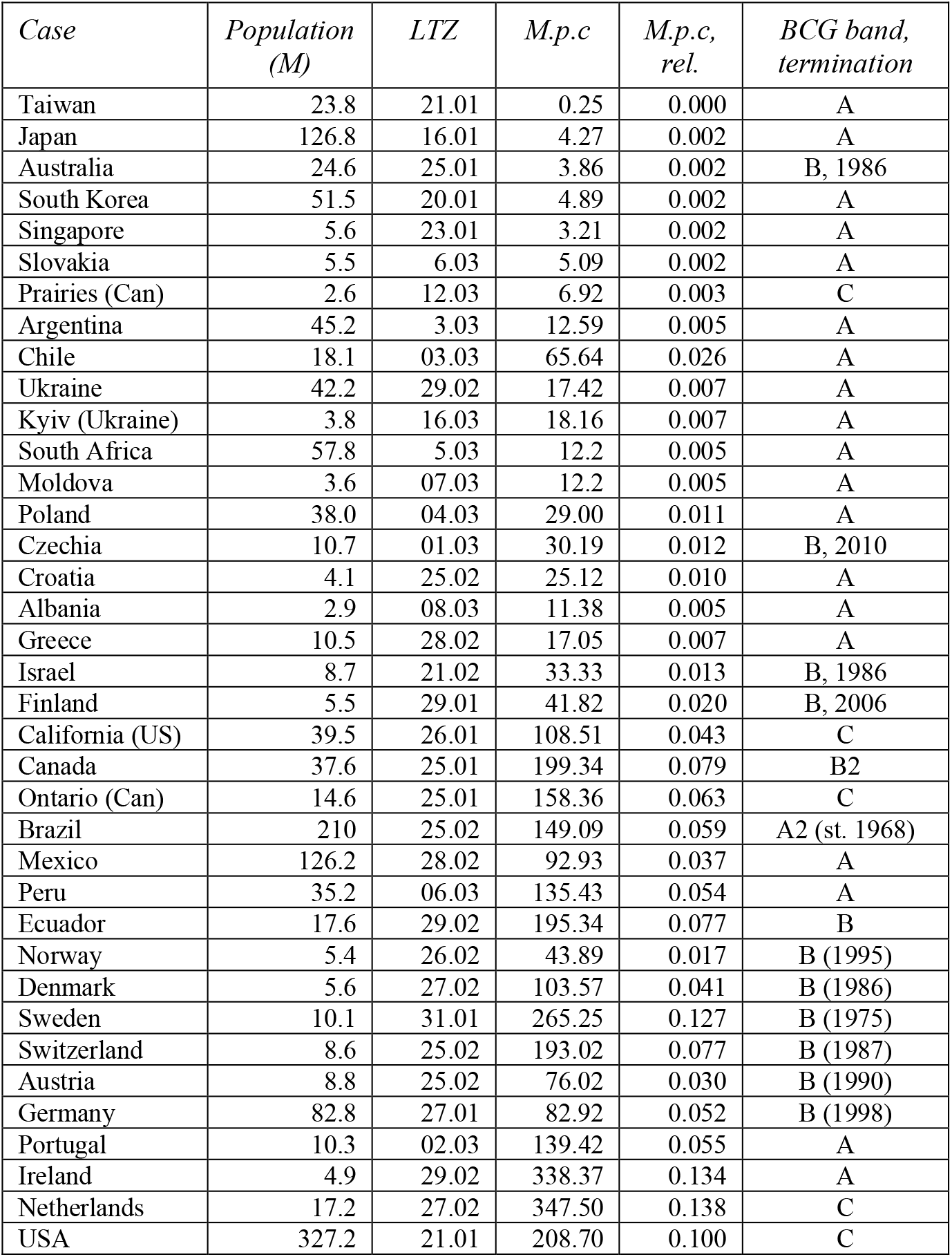
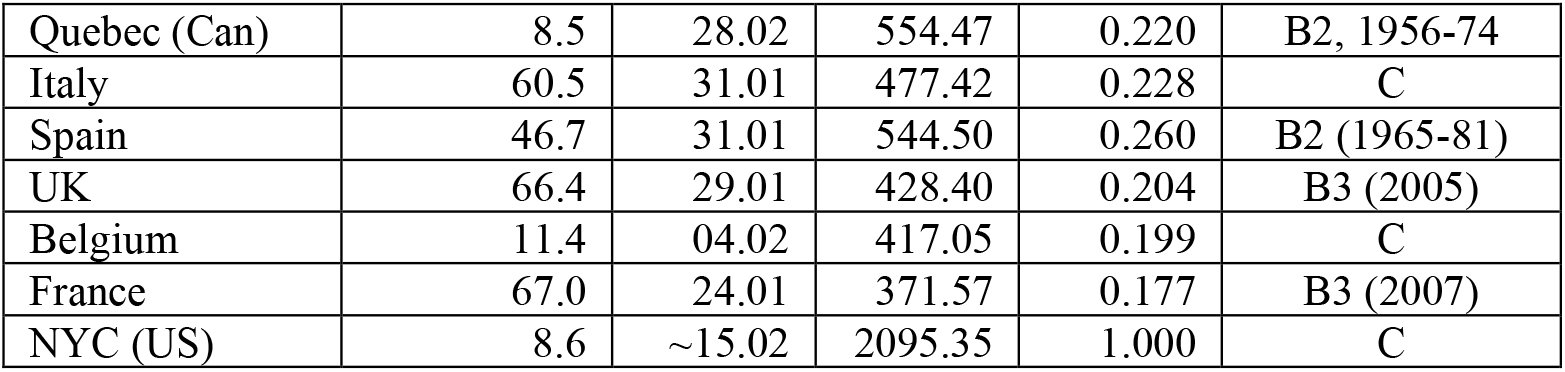
Time-Adjusted Combined Dataset, @LTZ + 3 m (updated 04.06.2020)

**Table 3.**
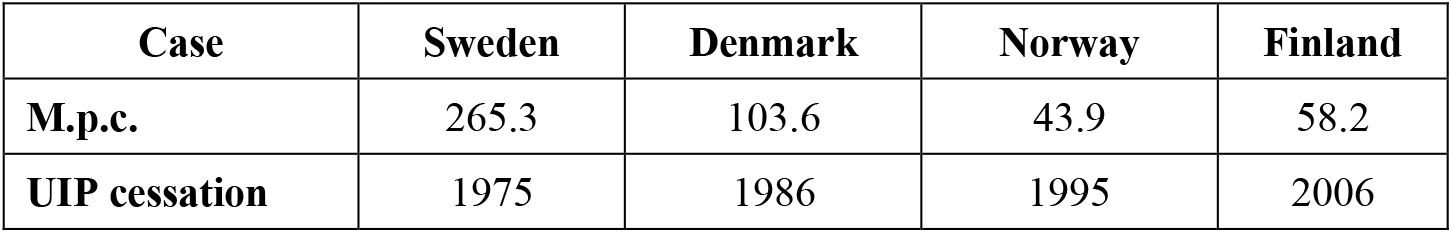
Covid-19 impact vs. Termination of BCG UIP, Northern Europe.

#### South America

All reviewed cases in South America with a current UIP show lower impact than those without it (Table 4).

**Table 4.** Covid-19 impact South America.

| Case | Peru | Mexico | Brazil | Ecuador |
| --- | --- | --- | --- | --- |
| **M.p.c.** | 135.4 | 92.9 | 149.1 | 195.3 |
| **Current UIP** | yes | yes | yes (late start) | no |

**Table 5.** Group samples.

| Sample | Size (number of cases) | Sample mean | Sample STD |
| --- | --- | --- | --- |
| A (and equivalent) | 18 | 4.42 | 2.42 |
| C (and equivalent) | 12 | 8.26 | 1.43 |

#### New York vs Kyiv

The cities offer almost a classical case for a comparison analysis with a very clear separation by most factors. Large and dense urban centers with high concentration of population, a large network of public transport, bustling social, commercial and entertainment hubs. Both are busy and popular hubs of national and international travel. Since 2017 Ukraine has a visa-free travel agreement with the European Union.

Kyiv, Ukraine: population 4 million; group A (current BCG UIP) New York City, USA: population 8.6 million; group C (no UIP)

The conclusions of the original analysis are strongly maintained at LTZ + 3 m: NYC, M.p.c. (03.05.2020): *2095* vs. Kyiv (03.06.2020): *18.2*.

### 2.6 Rapid Onset Cases in BCG Group A

Several cases of rapid onset of Covid-19 disease were reported in countries with a current BCG UIP, including but probably not limited to, the following: Iran; Russia; China; Brazil; Ireland. Without going into specific details of each case that can be done in another study, we will make some general observations.

On the assumption that a UIP is being offered from around 1950-ies (before that, the effects of WWII would likely compromise any such plans in the cases of significance) large groups of population would remain not covered even in the group A countries, such as age cohort above 70, people with chronic illness etc. Consequently, these groups would remain at a higher risk of a heavier outcome even in the scenario with induced immunity.

In cases where these vulnerable groups would happen to be more exposed to the infection, it is possible to see higher impact numbers, and rapid onset of the infection. At least three such scenarios can be named immediately: 1) a tradition of compact and large family dwelling with representatives of different age groups; 2) a practice of concentrated group residence of vulnerable people in a close community; 3) a record of prolonged social disorder that could have compromised the administration of UIP. Most of the observed rapid onset cases in group A fall into one of these categories, though certainly a more detailed analysis of those cases is warranted.

### 2.7 Possible Mechanism

The hypothesis of induced early age immunity protection from the exposure to BCG proposed in [2] based on a number of reports pointing at a possible association between early delivery of BCG vaccine and a broad immunity against several conditions [13-15]. It is further supported by a study indicating a possible mechanism for increased production of immune cells in infants following vaccination with BCG [16].

If boosting the production of immune cells can be confirmed to have a lasting effect, it could certainly contribute to the explanation of the epidemiological patterns discussed earlier that appear to indicate a correlation between universal immunization and a milder epidemiological scenario of Covid-19.

## 3 Statistical Significance of the Correlation Hypothesis

With the accumulated data of 40 cases in each of the time-adjusted datasets (LTZ+2m, LTZ+3m, plus Wave 1 dataset at LTZ+ 4m (early June, 2020)), approximately 100 data points overall, a question about statistical significance of the correlation hypothesis can be approached quantitatively. The analysis will be based on the distribution of the epidemiological impact measured by a logarithm of M.p.c. in the groups by BCG UIP record.

The null hypothesis would imply that immunization would carry no statistical significance for the epidemics impact, and therefore distributions in all of BCG group sample points (A, B, C) as defined above described by a single distribution with, in all likelihood, time-dependent parameters *μ(t), σ(t*) that can be estimated from the overall dataset distribution to have a standard deviation σof ∼2.55. The basis for the analysis that follows is the observation of a strong disparity between the sample means in groups A and C as seen in Fig.1, Section 2.3. Under the null hypothesis, whereby immunization has no significant effect on the impact, these cases should be treated as a difference between the means of randomly drawn samples of a given length, for which distribution parameters can be estimated with the sample-mean law.

Because all three samples (in each dataset) are drawn from the same null hypothesis distribution, the rule of sample means dictates that the means of the samples of groups A – C with the number of samples N_S_ will be distributed with the same mean and a standard deviation *σ*_*S*_ as:

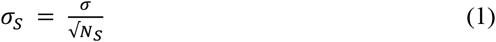

From (1) based on the count of points in each group, one can estimate sample mean standard deviations for the groups A and C samples, as, respectively, *0.6* and *0.74*.

To satisfy the null hypothesis, the means of samples A and C would need to, both and independently, satisfy the normal distribution laws with the same mean *μ*_*S*_ and *σ*_*S*_ defined by (1). Easy to see that the probability of the null hypothesis would be maximized if both of the sample means *μ*_*A*_ and *μ*_*C*_ were at the minimal distance from *μ*_*S*_ i.e. the latter positioned at the midpoint between *μ*_*A*_ and *μ*_*C*_, so that *μ*_*S*_ = (*μ*_*C*_ *– μ*_*A*_)/2.

With the mean of the sample means estimated, one can easily calculate the probability of the null hypothesis as:

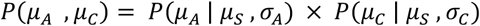

where the first term on the right is the probability of *μ*_*A*_ within the observed range below *μ*_*S*_ with a standard deviation *σ*_*A*_ and the second, similarly, of *μ*_*C*_ within the observed range above *μ*_*S*_ with a standard deviation *σ*_*C*_.

Then with the values of sample means and sample mean standard deviations obtained above the p-value of the null hypothesis can be estimated as:

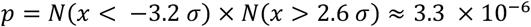

excluding the null hypothesis at a confidence level of at least 10^−5^.

Further, the distributions of BCG group samples in the time point datasets under the assumption of the null hypothesis must be independent between the datasets as well, so that accumulation of data can further improve the confidence of the correlation hypothesis.

To summarize the results of this analysis, if the immunization group samples had no correlation with the impact and therefore, considered as independent random samples under the null hypothesis, repeated observations of sample means as far apart as in the observations of BCG groups would lead to a strong estimate for the p-value of the null hypothesis and the resulting rejection conclusion. Note that for the sake of consistency, in group A were included the cases of Ireland and Brazil, both in the subgroup A2 with certain qualifications on UIP (Ireland: consistency of administration practice [17]; Brazil: late start of the immunization program). Exclusion of these cases would likely have resulted in a significantly lower sample mean and STD for this group and consequently, stronger constraints on the null hypothesis.

## 4 Conclusion

We hope that time-adjusted datasets compiled in this work as the early observations obtained with it can be useful to other researchers in the field looking for effective approaches to understanding and eventually, taking the pandemics under control.

In addition to convincing, in our view, arguments in favor of the BCG immunization correlation hypothesis presented in Sections 2.2 – 2.6, the statistical analysis of the correlation between BCG UIP and milder epidemiological scenarios confirms statistical significance of the correlation with confidence of at least 0.0001 providing a strong rationale for further studies that would investigate possible mechanism for such protection with the potential of developing effective methods of long-term immunity development against a broad range of diseases.

## Data Availability

Data referred to was publicly available online at the time of last access

http://www.bcgatlas.org/

https://www.google.com/covid19-map/

https://www.canada.ca/en/public-health/services/diseases/2019-novel-coronavirus-infection.html?topic=tilelink

https://www1.nyc.gov/site/doh/covid/covid-19-data.page

## Appendix Case Datasets

## References

1. Dolgikh S., Further evidence of a Possible Correlation Between the Severity of Covid-19 and BCG Immunization, MedRxiv, https://www.medrxiv.org/con-tent/10.1101/2020.04.07.20056994v1 April 20.

2. Zwerling A., Behr M.A., Verma A., Brewer T.F., Menzies D., Pai M. The BCG World Atlas: A Database of Global BCG Vaccination Policies and Practices, PLOS Medicine https://doi.org/10.1371/journal.pmed.1001012, 2011.

3. Covid-19 timeline ABC new https://abcnews.go.com/Health/timeline-corona-virusstarted/story?id=69435165 (14.03.2020)

4. BCG World Atlas http://www.bcgatlas.org/

5. Coronavirus map Google https://www.google.com/covid19-map/ (4.04.2020).

6. Canada Covid-19 situation update https://www.canada.ca/en/public-health/services/dis-eases/2019-novel-coronavirus-infection.html?topic=tilelink (4.04.2020)

7. Health Ministry of Ukraine https://www.kmu.gov.ua/en/news/v-ukrayini-zareyestrovanijvipa-dok-zahvoryuvannya-na-covid-19-moz (4.04.2020).

8. NYC Covid-19 Updates https://www1.nyc.gov/site/doh/covid/covid-19-data.page

9. Taiwan Center for Disease Control Covid-19 information https://www.cdc.gov.tw/En/Cate-gory/ListContent/bg0g_VU_Ysrgkes_KRUDgQ (30.03.2020).

10. Rousseau M.C., Conus F., Ka K. et al. Bacillus Calmette-Guérin (BCG) vaccination patterns in the province of Québec, Canada, 1956–1974, Vaccine 35 (36), 4777–4784, 2017.

11. Faust, L., Schreiber, Y., Bocking N. A systematic review of BCG vaccination policies among high-risk groups in low TB-burden countries: implications for vaccination strategy in Canadian indigenous communities. BMC Public Health 19, 1504, 2019.

12. Miller A., Reandelar M-J., Fasciglione K., Roumenova V., Li Y., Otazu G.H. Correlation between universal BCG vaccination policy and reduced morbidity and mortality for COVID-19: an epidemiological study, medRxiv 2020.03.24.20042937 2020.

13. Brandau S.; Suttmann H., Thirty years of BCG immunotherapy for non-muscle invasive bladder cancer: a success story with room for improvement. Biomed Pharmacother, 61 (6), 299–30 2007.

14. Roy P., Vekemans J., Clark A. et al. Potential effect of age of BCG vaccination on global paediatric tuberculosis mortality: a modelling study, The Lancet, 7(12), pe1655–1663, 2019.

15. Channappanavar R, Fett C, Mack M, Ten Eyck PP, Meyerholz DK, Perlman S. Sex-Based Differences in Susceptibility to Severe Acute Respiratory Syndrome Coronavirus Infection. Journal of Immunology, 198(10), 4046–4053, 2017.

16. B. Brook, D.J. Harbeson1, C.P. Shannon et al., BCG vaccination–induced emergency granulopoiesis provides rapid protection from neonatal sepsis, Science Translational Medicine. 12 (542), May 2020.

17. Sweeney E., Dahly D., Seddiq N., et al., Impact of BCG vaccination on incidence of tuberculosis disease in southern Ireland, BMC Infectious Diseases, 19. 397, 2019.

